# Cortico-Limbic and Sensorimotor Network Connectivity Link Brain Function to Atherosclerosis in Chronic Stress: Beyond Amygdala-PFC

**DOI:** 10.1101/2025.08.29.25334386

**Authors:** David O’Connor, Mandy M.T. van Leent, Philip M. Robson, Shady Abohashem, Charbel Gharios, Audrey E. Kaufman, Michael T. Osborne, Maria Giovanna Trivieri, James W. Murrough, Lisa M. Shin, Ahmed Tawakol, Zahi A. Fayad

**Author notes:** **Addresses for correspondence:** Ahmed Tawakol, MD, Co-director, Cardiovascular Imaging Research Center, Associate Professor of Medicine, Cardiology Division, Massachusetts General Hospital and Harvard Medical School, 55 Fruit St, Yawkey 5E, Boston, MA 02114-2750, USA, Twitter: @ATawakolMD; Zahi A. Fayad, PhD, Lucy G. Moses Professor in Medical Imaging and Bioengineering, Vice Chair for Research and Professor, Department of Diagnostic, Molecular and Interventional Radiology, Professor, Department of Medicine, Division of Cardiology, Director, Biomedical Engineering and Imaging Institute, Icahn School of Medicine at Mount Sinai, One Gustave L. Levy Place, Box 1234 New York, NY 10029-6574, USA, Twitter: @zahifayad. These authors contributed equally (Ahmed Tawakol, Zahi A. Fayad).

## Abstract

Chronic stress is a recognized risk factor for atherosclerotic cardiovascular disease (CVD), yet the underlying biological mechanisms remain incompletely understood. Dysregulation of cortico-limbic brain circuits, leading to heightened systemic inflammation and accelerated progression of CVD risk factors, has been proposed as a central pathway. Prior research has primarily focused on altered connectivity between the amygdala and the prefrontal cortex (PFC); however, the contributions of additional cortical and subcortical regions have not been fully delineated.

In this study, we utilized a multimodal imaging approach, integrating brain magnetic resonance imaging (MRI) and vascular imaging, to assess both functional and structural connectivity between the amygdala and broader brain networks. Consistent with previous findings demonstrating increased inflammation, greater atherosclerotic burden, and impaired amygdala– PFC connectivity in chronically stressed individuals, we show that task-based functional and structural connectivity measures independently distinguish participants with higher versus lower atherosclerotic burden. Importantly, while limbic and prefrontal regions remain critical, our findings also highlight brain regions involved in sensorimotor and autonomic processes, including the sensorimotor cortices and cerebellum, in the stress–atherosclerosis pathway.

By expanding the scope beyond the amygdala–PFC axis, these results offer a more comprehensive framework for understanding the neural mechanisms linking chronic stress to CVD and may guide the development of novel therapeutic strategies aimed at neuroimmune modulation.

## Introduction

### Chronic stress and Cardiovascular Disease

Chronic stress is associated with increased levels of cardiovascular disease (CVD) ^1,2^. The mechanism underlying this relationship is still incompletely understood ^3^. Prior work has suggested that an imbalance in cortico-limbic brain function is a key part of this process. The amygdala – a critical region within the limbic system - forms a central part of this mechanism, with aberrant amygdala activity leading to more frequent activation of the sympathetic nervous system upregulation of inflammation, and stimulation of the hypothalamic-pituitary-adrenal (HPA) axis ^4^. The associate effects of frequent stimulation of the HPA axis include consistently elevated levels of stress hormones and inflammatory markers in the blood, such as C-reactive protein (CRP) and interleukin-6 (IL-6), which are associated with vascular inflammation and atherosclerotic plaque formation ^5^.

Stress-CVD related investigations have focused largely on the relationship between the amygdala and prefrontal cortex (PFC). This construct obscures the role of other cortical regions, in particular sensorimotor cortices and the cerebellum. These regions may be particularly relevant given their role in autonomic function ^6–9^. Findings in murine models further support this perspective; in contrast to PFC activity, which dampens limbic-driven stress responses, heightened activity within the motor cortex induces rapid neutrophil mobilization from the bone marrow to peripheral tissues through skeletal muscle-derived neutrophil-attracting chemokines ^10^. These data demonstrate that distinct brain regions differentially and rapidly tailor the leukocyte landscape during psychological stress, suggesting that sensorimotor areas may play a more active role in neuroimmune regulation than previously appreciated. A more wholistic approach to brain connectivity may provide greater insight into cortico-limbic dysregulation and its association with chronic stress disorders.

### Chronic Stress, Trauma Exposure, and PTSD

Chronic stress results from prolonged exposure to repetitive stressful stimuli, leading to both physiological and psychological strain. Although stress responses are adaptative in the short term, chronic stress activates a cascade of neuroendocrine, immunologic, and autonomic responses that become maladaptive when sustained, resulting in systemic dysregulation and increased allostatic load ^11^. Similar biological disruptions are observed across a range of non-traumatic stress exposures such as high-pressure work environments and social isolation; however, a more severe manifestation can result from trauma involving threatened death or serious injury such as in post-traumatic stress disorder (PTSD). PTSD illustrates the profound consequences of prolonged stress activation and is characterized by intrusive, distressing memories and emotional dysregulation, often accompanied by an imbalance in cortico-limbic brain function ^12^. Beyond neural disruptions, PTSD is associated with heightened inflammation ^13^, and individuals with PTSD have higher levels of atherosclerotic burden, coronary artery disease, and elevated risk of CVD events ^14–16^. These findings suggest that PTSD serves as an effective model for exploring the mechanisms linking chronic stress to cardiovascular disease^17,18^.

### Imaging Studies

Multimodal multi-organ imaging facilitates investigating brain and cardiovascular health simultaneously ^19,20^. Magnetic resonance imaging (MRI), in particular functional and diffusion MRI, has been widely used to assess brain function and structure in health and disease ^21–23^. Structural MRI of the vasculature has also been used to quantify plaque burden, including measurements of vessel wall thickness in subclinical disease ^24–27^. In recent studies, fluorodeoxyglucose – positron emission tomography (FDG-PET) has been established as an effective tool for assessing stress related brain metabolic activity as determined by uptake of FDG, a radiotracer analogue of glucose ^19,28^. Given the temporal variance of physiological processes relating to stress and brain function, simultaneous acquisition of MRI and PET is essential for investigating integrated biological systems. The complementary strengths of these modalities support the use of hybrid PET-MR systems for comprehensive cardiovascular and brain imaging.

Neuroimaging has been used extensively in stress investigation ^29^, and in PTSD in particular ^30–33^. The limbic system, which is heavily involved in emotional regulation, reoccurs in findings, in particular the structure and function of the amygdala and hippocampus and their connectivity with the PFC. Other regions have also been implicated, for example association cortices and the cerebellum, and how they integrate sensorimotor information are thought to also play a role ^34–37^. Acquiring detailed multimodal imaging of brain structure and function alongside structural and molecular imaging of vasculature can shed light on the relationship between chronic stress and CVD. Prior work from our group has shown increased systemic inflammation and higher atherosclerotic burden in chronically stressed individuals ^38^, and underscored the importance of the amygdala-PFC connection. The mechanisms leading to dysregulated cortico-limbic function and structure are unclear but will naturally involve brain networks beyond the limbic system and PFC.

### Our study

The causal relationship between chronic stress and cardiovascular disease (CVD) remains difficult to establish due to the complex interplay of biological, psychological, and social factors ^3^. To further elucidate underlying mechanisms, we investigated whether MRI-based functional and structural connectivity between the amygdala and cortical, subcortical, and cerebellar regions could stratify participants by atherosclerotic burden, assessed through carotid vessel wall MRI, across a population with varying levels of chronic stress.

## Results

### Dataset grouping

Our initial goal was to determine if functional and structural connectivity in the brain could predict CVD burden in participants with PTSD, trauma controls and healthy controls. We focused on network level connectivity with the amygdala, as prior research has underscored its importance ^28,38^. Participants’ data were regrouped based on cardiovascular health, specifically using a median split on atherosclerotic plaque burden in the bilateral carotid arteries, as assessed by standard deviation of the vessel wall thickness (SDWT-C). This resulted in two groups, a higher burden group, and a lower burden group. The median burden and grouping varied slightly across analyses and depended on availability of good quality task fMRI (t-fMRI), resting state functional MRI (rs-fMRI), and diffusion MRI (dMRI), to ensure an equal (or close to equal) number of participants in the higher and lower groups. These group characteristics, split by modality, are shown in **Table 1**.

**Table 1.** Characteristics of the atherosclerotic burden-based subgroups. Listed variables are number of participants (N), trauma subgroups (PTSD: post-traumatic stress disorder, TC: trauma control, HC: healthy control), age, sex, in scanner participant motion (mean framewise displacement, mean FD), atherosclerotic burden (standard deviation of wall thickness of the bilateral carotids, SDWT Carotid), tobacco use (“Not at all / Never”, “Less than monthly”, “Monthly”, “Weekly”, “Daily or almost daily”), use of hypertension medication (yes, no), high blood pressure (never, past, current), and diabetes (yes, no). Values for continuous variables listed are “mean (standard deviation)”.

| t-fMRI | All | Lower Atherosclerosis Burden | Higher Atherosclerosis Burden |
| --- | --- | --- | --- |
| N | 60 | 30 | 30 |
| Trauma Subgroups | PTSD: 17, TC: 28, HC: 15 | PTSD: 7, TC: 13, HC: 10 | PTSD: 10, TC: 15, HC: 5 |
| Age | 39.37 (9.19) | 37.93 (8.86) | 40.80 (9.44) |
| Sex | Male: 23, Female: 37 | Male: 8, Female: 22 | Male: 15, Female: 15 |
| In scanner motion (mm) | 0.15 (0.04) | 0.15 (0.05) | 0.15 (0.04) |
| Atherosclerotic Burden | 0.14 (0.02) | 0.12 (0.01) | 0.16 (0.02) |
| Tobacco Use | Not at all / Never: 57,<br>Less than monthly: 1,<br>Daily or almost daily: 2 | Not at all / Never: 28,<br>Daily or almost daily: 2 | Not at all / Never: 29,<br>Less than monthly: 1 |
| Hypertension Meds | No: 56,<br>Yes: 4 | No: 30 | No: 26,<br>Yes: 4 |
| High Blood Pressure | Never: 57,<br>Past: 2,<br>Current: 1 | Never: 29,<br>Past: 1 | Never: 28,<br>Past: 1,<br>Current: 1 |
| Diabetes | No: 60 | No: 30 | No: 30 |
| rs-fMRI |  |  |  |
| N | 55 | 28 | 27 |
| Trauma Subgroups | PTSD: 15, TC: 23, HC: 17 | PTSD: 8, TC: 11, HC: 9 | PTSD: 7, TC: 12, HC: 8 |
| Age | 37.98 (8.56) | 37.18 (8.44) | 38.81 (8.76) |
| Sex | Male: 18, Female: 37 | Male: 7, Female: 21 | Male: 11, Female: 16 |
| In scanner motion (mm) | 0.16 (0.05) | 0.16 (0.06) | 0.16 (0.04) |
| Atherosclerotic Burden | 0.14 (0.02) | 0.12 (0.01) | 0.16 (0.02) |
| Tobacco Use | Not at all / Never: 54,<br>Daily or almost daily: 1 | Not at all / Never: 27,<br>Daily or almost daily: 1 | Not at all / Never: 27 |
| Hypertension Meds | No: 52,<br>Yes: 3 | No: 28 | No: 24,<br>Yes: 3 |
| High Blood Pressure | Never: 53,<br>Past: 1,<br>Current: 1 | Never: 28 | Never: 25,<br>Past: 1,<br>Current: 1 |
| Diabetes | No: 55 | No: 28 | No: 27 |

*Table 1- Characteristics of the atherosclerotic burden-based subgroups.
| dMRI |  |  |  |
| --- | --- | --- | --- |
| N | 65 | 33 | 32 |
| Trauma Subgroups | PTSD: 18, TC: 32, HC: 15 | PTSD: 9, TC: 14, HC: 10 | PTSD: 9, TC: 18, HC: 5 |
| Age | 39.32 (9.14) | 37.33 (8.43) | 41.38 (9.51) |
| Sex | Male: 29, Female: 36 | Male: 12, Female: 21 | Male: 17, Female: 15 |
| In scanner motion (mm) | 0.54 (0.13) | 0.54 (0.13) | 0.55 (0.14) |
| Atherosclerotic Burden | 0.14 (0.03) | 0.12 (0.01) | 0.16 (0.2) |
| Tobacco Use | Not at all / Never: 60,<br>Less than monthly: 2,<br>Daily or almost daily: 3 | Not at all / Never: 30,<br>Less than monthly: 1,<br>Daily or almost daily: 2 | Not at all / Never: 30,<br>Less than monthly: 1,<br>Daily or almost daily: 1 |
| Hypertension Medication | No: 60,<br>Yes: 5 | No: 32, Yes: 1 | No: 28, Yes: 4 |
| High Blood Pressure | Never: 61,<br>Past: 2,<br>Current: 2 | Never: 32,<br>Past: 1 | Never: 29,<br>Past: 1,<br>Current: 2 |
| Diabetes | No: 65 | No: 33 | No: 32 |

### Classification results

#### fMRI

Network level task-based connectivity between the right amygdala and whole brain was able to classify individuals with higher and lower plaque burden (70% Accuracy, p=0.011, ROC-AUC=0.75). In contrast, classification using left amygdala connections and bilateral connections were not successful. These results are shown in **Table 2**, first row. Classification based on covariates of non-interest only, including age, sex and scanner motion, performed worse than the right amygdala task-based model (61.7% Accuracy, p=0.085, ROC-AUC=0.56). The permutation test results and ROC-AUC for the successful t-fMRI model are graphed in **Supplemental Figure 1, Panel A**. No rs-fMRI model was successful, **Table 2**, second row.

**Table 2.** Results of classification modelling for predicting atherosclerotic burden. Variables of non-interest include age, sex, and participant motion in scanner.

| *Prediction of atherosclerotic burden* | Left Amygdala | Right Amygdala | Bilateral Amygdala | Variables of non-interest |
| --- | --- | --- | --- | --- |
|  | Accuracy (p value) | Accuracy (p value) | Accuracy (p value) | Accuracy (p value) |
| t-fMRI | 56.7% (p = 0.247) | **70.0% (p = 0.011)** | 61.7% (p = 0.091) | 61.7% (p = 0.085) |
| rs-fMRI | 49.3% (p = 0.534) | 50.7% (p = 0.488) | 45.0% (p = 0.722) | 47% (p = 0.636) |
| dMRI | **72.1% (p = 0.004)** | 49.3% (p = 0.525) | 61.2% (p = 0.069) | 52.9% (p = 0.397) |

#### Diffusion MRI

Structural connectivity between the left amygdala and whole brain was able to classify individuals with higher and lower plaque burden (72.1% Accuracy, p=0.004, ROC-AUC=0.73). Classifications using right amygdala connections and the average of the bilateral connections was not successful (**Table 2**). Classification of plaque burden based on age, sex and scanner motion performed worse than the left amygdala structural model (52.9% Accuracy, p=0.397, ROC-AUC=0.56). The permutation test results, and ROC-AUC, for the successful dMRI model are graphed in **Supplemental Figure 1, Panel B**.

### Model interpretation

The model classification performance was based on tenfold cross validation (CV). This allows assessment of how model weights vary based on subsamples of the dataset. Naturally, model weights which exhibit consistent magnitude and sign are of particular interest. Model weights for each successful model are shown in **Figure 1** and **Figure 2**, respectively. In each figure panel A shows the distribution of model weights across folds with a plus or minus symbol next to y axis label indicating the weight distributions which do not change sign across folds. Model weights that change sign across folds indicate that the corresponding predictive/independent variable, in this case, a connectivity value, could not be reliably associated with the target/dependent variable. For the task fMRI model in **Figure 1**, increased bilateral connections between the frontoparietal network (encompassing the ventromedial PFC, vmPFC) and the amygdala were associated with decreased CVD burden, in particular, the ipsilateral connection, a finding that is consistent with prior work ^38^. Notably, however, higher connectivity of bilateral medial frontal and motor cortices with the right amygdala corresponded with <u>higher</u> CVD burden. The subcortical, default mode, and visual/visual association networks showed strong hemispheric biases.

**Figure 1.**
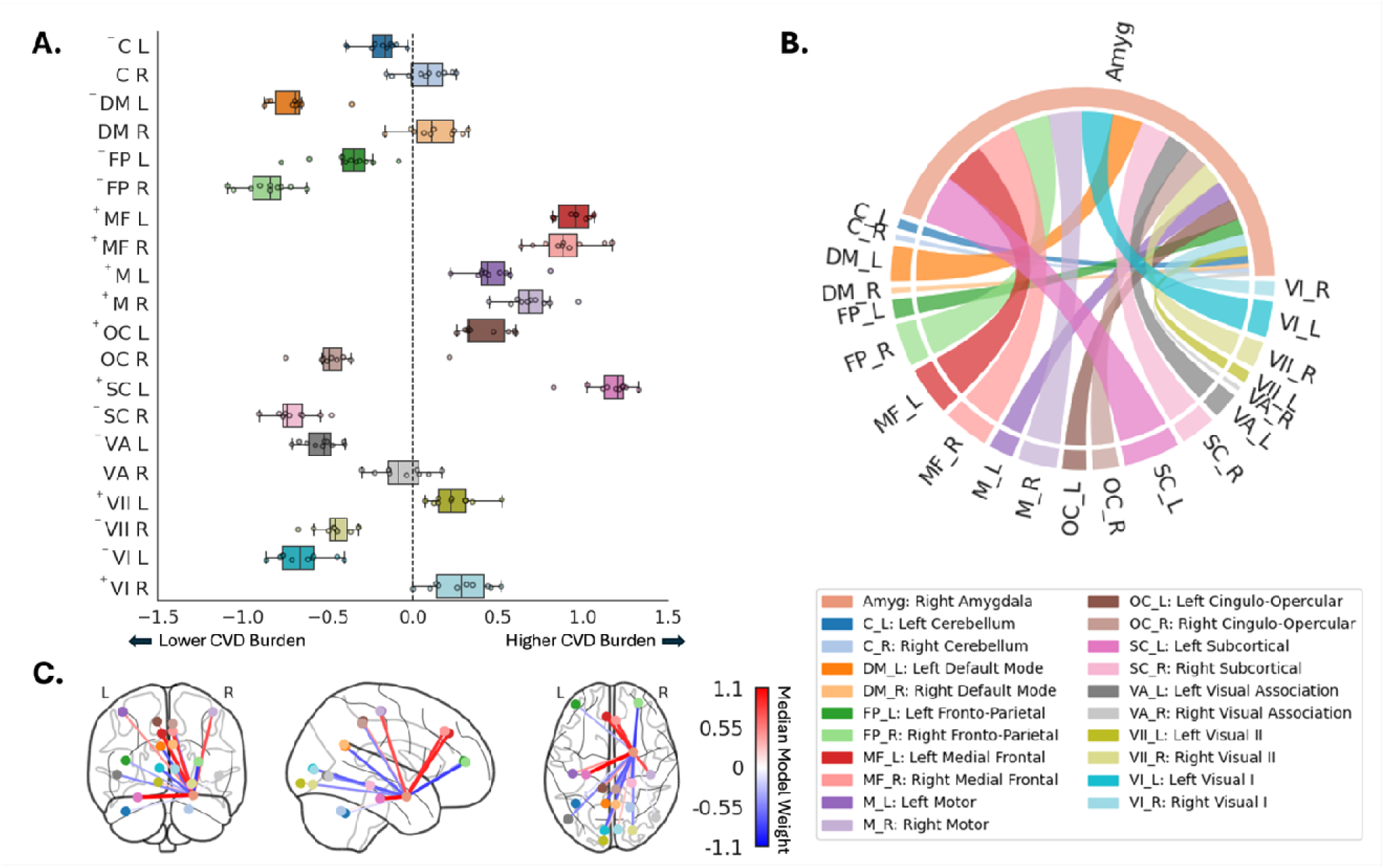
Model weights for the right amygdala-based task fMRI model. A) A box and whiskers plot of the logistic regression model weights across cross validation folds. These weights correspond to how much influence a feature, in this case a functional connectivity value, had on the model output. The midline of the box represents the median weight, the left and right sides of the box represent the 25th and 75^th^ percentile of the weights respectively, and the left and right whiskers represent 1.5 times the interquartile range from the 25^th^ and 75^th^ percentiles respectively. A plus next to a label on the y axis signifies that all model weights for a given network are above zero, and a minus all below zero. These symbols indicate that the relationship did not change direction across folds. B) A chord plot of the absolute value of the median model weight. C) A ball and stick plot also reflecting the median model weights. The “ball” reflects the centroid of a given network. The color of the boxes, chords and balls are consistently colored based on network and hemisphere combinations reflecting the underlying connectivity. Legend is show bottom right.

**Figure 2.**
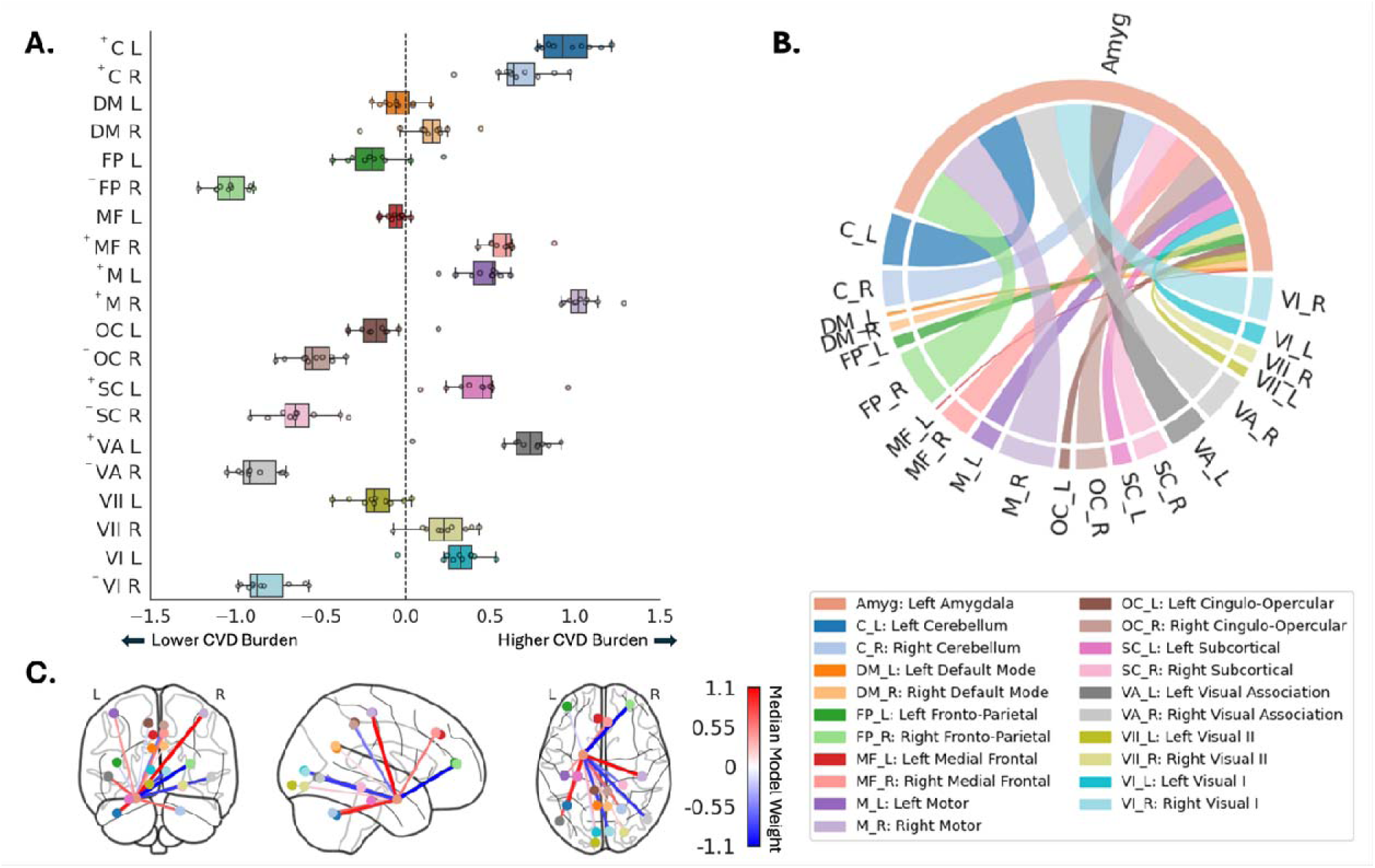
Model weights for the left amygdala-based dMRI model. A) A box and whiskers plot of the model weights across cross validation folds. . These weights correspond to how much influence a feature, in this case a structural connectivity value, had on the model output. The midline of the box represents the median weight, the left and right sides of the box represent the 25th and 75^th^ percentile of the weights respectively, and the left and right whiskers represent 1.5 times the interquartile range from the 25^th^ and 75^th^ percentiles respectively. A plus next to a label on the y axis signifies that all model weights for a given network are above zero, and a minus all below zero. These symbols indicate that the relationship did not change direction across folds. B) A chord plot of the absolute value of the median model weight. C) A ball and stick plot also reflecting the median model weights. The “ball” reflects the centroid of a given network. The color of the boxes, chords and balls are consistently colored based o network and hemisphere combinations reflecting the underlying connectivity. Legend is shown bottom right.

For the dMRI model in **Figure 2**, bilateral motor cortices also contributed to the model in that higher connectivity with the left amygdala corresponded with higher CVD burden. Again, increased bilateral connections between the frontoparietal network, and the amygdala were associated with decreased CVD burden. Once more the subcortical, default mode, and visual/visual association networks showed hemispheric biases, though the default mode weight distributions overlapped with zero, and thus the connections with the amygdala are negligible in this context.

In both modalities, increased connectivity with the frontoparietal network is associated with lower atherosclerosis burden, while increased connectivity with the motor cortex is associated with higher atherosclerosis burden. Visual cortices showed distinct opposite patterns in each modality due to hemispheric bias.

### Composite network score

We generated composite scores for each of the successful task and diffusion models. This was done by calculating the dot product of the median model weights across CV folds, for the median performing model, and the underlying connectivity estimates for each participant. This yielded one composite score per image, per participant. These composite scores represent a CVD sensitive brain connectivity metric, which can then be assessed for its association with other variables of interest. For each of the successful MRI-based models the composite scores significantly associated with SNA (task fMRI; β = 0.238, p value = 0.003, dMRI; β = 0.162, p value = 0.007), as well as systemic inflammation (task fMRI; β = 0.366, p value = 0.007, dMRI; β = 0.243, p value = 0.047). These associations were estimated using OLS, correcting for age, sex and in-scanner motion, and are depicted in **Supplemental Figure 2**. This suggests that the brain connectivity networks that were successful in classifying higher and lower atherosclerotic burden are also associated with systemic inflammation and SNA. However, the composite scores did not associate with self-reported stress and resilience, structural assessment of the aorta, FDG uptake in the carotids and aorta, nor inflammation as assessed by FDG-PET of the bone marrow and spleen. A table of all results in shown the “Regression Results” section of the Supplemental Information.

### PET co-registration & new SNA values & associations

Given that classification results suggest that regions beyond the PFC impact the amygdala and may also be useful for assessing CVD, we explored whether normalizing amygdalar FDG uptake by other brain regions could be relevant. Using the fMRI-based Shen 368 atlas ^39^, we normalized amygdalar FDG uptake by each atlas-defined region and performed OLS regression to assess associations with atherosclerotic burden (SDWT-C). **Figure 3** shows the results, highlighting significant associations (uncorrected for multiple comparisons) when normalizing by motor, visual, and cerebellar regions. In all cases, adjusted t-statistics from an OLS model (controlling for age and sex) were positive, indicating that higher relative amygdalar FDG uptake is associated with increased atherosclerotic burden.

**Figure 3.**
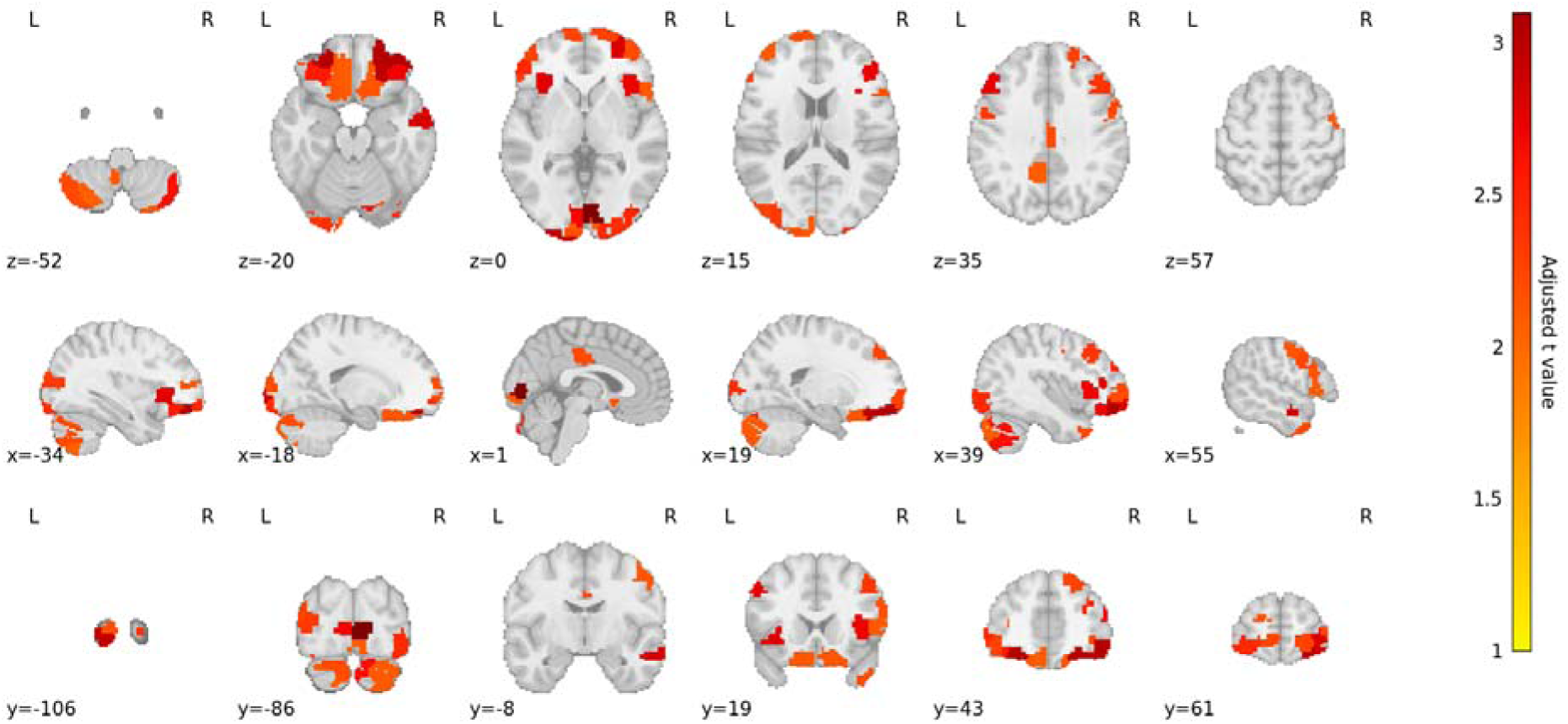
Brain regions where amygdala activity ratios associate with plaque burden. Shown are the adjusted t values controlling for age and sex. All t values were positive, i.e. increasing ratio values were associated with increased atherosclerosis. Each region represents the mean bilateral amygdala signal normalized by the mean signal of that region. Regions were defined by the Shen 368 atlas. Multiple comparison correction was not performed.

Each region in the analysis is associated with one of the networks used in the functional and structural connectivity analyses, allowing us to assess which networks were most represented in the PET analysis. A breakdown of the network membership of the regions in **Figure 3** is shown in **Supplemental Table 2.** The frontoparietal network, which includes the vmPFC, was the most represented network, aligning with prior findings ^28,38^. However, regions from across the brain, including motor, visual, medial frontal, default mode, and cerebellar regions, were also represented, with sensorimotor regions in aggregate (motor, visual I and II, and visual association) the second most represented group of brain regions.

## Discussion

Our findings highlight the heterogeneity of network-level amygdala connectivity associated with CVD burden, with distinct contributions from both functional and structural MRI measures. Task-based functional connectivity with the right amygdala and structural connectivity with the left amygdala effectively classified individuals with high versus low plaque burden, outperforming models based solely on demographic and motion-related covariates. Notably, increased frontoparietal network connectivity was associated with lower CVD burden, whereas increased motor cortex connectivity was associated with higher burden, underscoring the complexity of neural pathways linking stress to vascular health. Complementary PET imaging analyses supported these observations, demonstrating that normalization of amygdalar FDG uptake by FDG uptake in sensorimotor and cerebellar regions yielded novel metabolic imaging markers sensitive to CVD burden.

### Lateralization of amygdala

A consistent, but underappreciated, feature of the amygdalae is a significant hemispheric asymmetry in structure and function ^40^. The amygdalae show hemispheric asymmetries in volume ^41^, structural connectivity ^42–45^, functional activation ^46,47^ and functional connectivity ^48,49^. Crucially for this study, these differences are reflected in how the amygdalae react to emotional stimuli, including fear, pain processing and stress ^50–53^. It is thought that the right amygdala is more active in intermittently responding to emotional stimuli, while the left amygdala is more involved in sustained evaluation of stimuli ^54,55^. The differential response of the amygdalae to emotional stimuli may explain why cardiovascular burden is more associated with the left amygdala in our structural connectivity model, as this likely reflects a steady state of neuro-configuration, as opposed to the right amygdala in the task-based model, which reflects the dynamic processing of periodic emotional stimuli.

### Involvement of amygdala in motor, visual, cerebellar circuits

Beyond amygdala lateralization there is further heterogeneity in the function of amygdala nuclei, which is likely crucial to understanding a wide range of pathologies ^56^. Sylvester et al. have identified three distinct amygdala subdivisions with unique functional network associations, all of which tended to correlate with somatomotor networks. Structural evidence on an amygdala-motor pathway has also been shown using tractography in healthy adults ^57^.

Amygdala-motor interactions are further supported by research on conversion disorder, where the left amygdala shows increased functional connectivity with an inhibitory motor circuit during a sensorimotor/emotional stimulation task ^58^. In PTSD, increased resting state connectivity between anterior vermis, a cerebellar region associated with locomotion, and amygdala has been observed ^9^, with this connectivity linked to decreased heart rate variability. Further work shows cerebellar resting state connectivity differences in both healthy controls for fear conditioning, and in PTSD ^59–62^.

Recent findings indicate that the visual cortices responsible for more complex integrative visual processing can modulate subcortical regions in the context of fear, specifically fear suppression ^63^. This contrasts with past studies showing a “top-down” control of visual responses by higher level brain regions in subcortex, such as the amygdala ^64^. These findings may be reconciled by considering work showing a bidirectional communication between the fusiform gyrus and the amygdala has been shown ^65,66^. The relationship between the amygdala and visual cortex appears complex, with evidence suggesting that the visual cortex may play a larger role in modulating amygdala responses to emotional stimuli than previously thought.

Together these findings highlight the integration of subcortical regions with unimodal cortices and the cerebellum in the context of emotional processing. Further exploration of the interactions between unimodal cortical regions and the autonomic system could provide insight into the neural mechanisms underlying brain connectivity and CVD.

### Brain Connectivity and CVD

The findings in this study align with growing evidence underscoring the relevance of the brain-cardiovascular axis in CVD pathogenesis. Prior work has consistently highlighted the amygdala’s central role ^28,38^, demonstrating how neural activity patterns related to emotional processing and stress influence cardiovascular outcomes ^67,68^. Chronic stress can lead to synaptic remodeling in the amygdala ^69^, and reduced amygdala and hippocampal volumes ^70,71^, impacting the HPA axis ^4^, and elevating stress-related CVD risk. Functionally, the amygdala has been shown to modulate autonomic responses like heart rate variability ^72^, and stressor-evoked blood pressure reactivity through its connectivity with the pregenual anterior cingulate cortex, the latter correlating with vascular pathology ^73^. Furthermore, the amygdala-dorsomedial PFC pathway’s association with enhanced inflammatory responses ^74^, provides a crucial link between neural circuits and systemic inflammation, a known driver of CVD ^75^. The correlations between systemic inflammatory markers and amygdala-centered brain connectivity underscores the intricate role of the brain in mediating CVD.

Despite the substantial evidence linking amygdala function and structure with CVD, the mechanisms underlying abnormal amygdala function remain less clear. Beyond the amygdala, sensorimotor processes may play a significant role in shaping stress responses relevant to CVD. Research indicates that both motor and fear circuits contribute to regulating peripheral immune activity (e.g., leukocyte activity) during acute stress ^10^. Given the strong connections between sensorimotor circuits and autonomic regulation ^6,8^, and evidence of their dysfunction in stress-related disorders like PTSD ^9^, disruptions in these pathways could represent an important, potentially underappreciated, contributor to altered cardiovascular risk.

### Clinical relevance

Non-pharmacological interventions for chronic stress disorders, beyond cognitive behavioral therapy, have incorporated sensory and motor components. Eye movement desensitization and reprocessing (EMDR) uses guided eye movements, tapping, and/or auditory cues to reprocess traumatic memories, and has demonstrated effectiveness in alleviating PTSD symptoms ^76,77^. As well as alleviating symptoms, EMDR was associated with an increase in grey matter density in several areas of the PFC, including the inferior and middle frontal gyri, the medial PFC, and the anterior cingulate cortex ^76^, as well as reduced metabolic connectivity between the precuneus and cerebellum occurring after treatment ^77^.

Motor interference therapy (MIT) has also been shown to reduce trauma re-experiencing, anxiety and PTSD symptoms ^78,79^. MIT works on a similar basis to EMDR in which a patient mentally retains a traumatic memory or distressing though while being guided by an audio track directing them to complete simple motor and cognitive tasks. Visuospatial tasks can both distract the participant and interfere with sensory aspects of intrusive memories during trauma recall ^80,81^. It has also been shown that neuromodulation of the visual cortex using repetitive transcranial stimulation can reduce the intensity of intrusive memories in healthy individuals ^82^. The work described above highlights the role of sensory processing in managing trauma-related cognition.

Mindfulness training has also been recognized for managing chronic stress ^83^, improving PTSD symptoms ^84^, and changing how the amygdala responds to negative facial expressions ^85^. Mindfulness has also been shown to alter stress-related amygdala resting state functional connectivity ^86^ and increase connectivity within visual and auditory networks, which may reflect enhanced sensory processing ^87^. Crucially, mindfulness has proven useful in reducing hypertension ^88,89^, and individuals who received mindfulness training for reducing blood pressure saw increased axonal integrity in the fornix (a key node in the limbic system) along with decreased axonal integrity in the corticospinal tract and increased axonal integrity in the right cerebellar peduncle ^90^. The convergence of findings, highlighting the involvement of sensory processing, motor systems, and the cerebellum in effective stress/trauma therapies, alongside similar neural correlates in interventions reducing cardiovascular risk factors, suggests these mechanisms are pertinent to the brain-heart axis and warrant further investigation.

## Conclusions

We identified functional and structural brain connectivity networks that classified participants according to atherosclerotic plaque burden across a range of chronic stress levels. These networks were associated with stress-related neural metabolic activity and systemic inflammation. Our results reinforce the established link between amygdala–frontoparietal connectivity and stress-induced CVD, while also implicating the motor and visual cortices and the cerebellum as additional contributors. FDG PET analysis further demonstrated that differential metabolic activity between the amygdala and the motor, sensory, and cerebellar cortices was associated with vascular wall thickness, corroborating our MRI-based connectivity findings and suggesting broader sensorimotor and cerebellar involvement in stress-related CVD pathophysiology.

## Materials and Methods

### Dataset

Participants took part in the prospective study “PET/MRI of the Brain-Hematopoiesis-Atherosclerosis Axis in PTSD Patients” (ClinicalTrials.gov Identifier: NCT03279393). The dataset is described in detail by Gharios et al ^38^. Briefly, 94 participants completed the study and met inclusion criteria. Further criteria we applied for a given analysis depending on the availability and quality of the imaging data (See Table 1 for the number of participants used in each analysis). More details can be found in the Supplemental Information.

### Imaging

#### Overall Protocol

Subjects underwent whole body FDG PET and MRI imaging of the head, neck, chest, and abdomen. Injection of ∼10 mCi of FDG occurred 30 minutes before brain PET imaging started, and 90 minutes before vascular PET imaging started. MRI-based attenuation correction for PET data was performed standard two-point DIXON 3D gradient echo was used for MRAC as supplied by the vendor. A 3D T1-weighted MPRAGE sequence was used to acquire a high-resolution anatomical scan of the brain. More details can be found in the Supplemental Information.

#### fMRI

Both rs-fMRI and t-fMRI data were acquired. fMRI processing was performed using the fMRIPrep workflow ^91^. In short fMRI data were corrected for motion, susceptibility artifact, and physiological artifacts. fMRI data were then normalized to MRI space. fMRIPrep utilized FSL ^92^ and ANTs ^93^ as part of this processing. More details can be found in the Supplemental Information.

#### dMRI

Two opposite phase-encoded (AP, PA) acquisitions using a NODDI sequence were obtained for the diffusion weighted MRI of the brain. More details can be found in the Supplemental Information.

#### Vascular MRI

3D dark-blood MRI of the internal carotid arteries and ascending aorta was performed using a 3D SPACE sequence. More details can be found in the Supplemental Information.

#### Brain, Vascular and Leukopoietic PET

Brain function, leukopoietic activity in the bone marrow and spleen, and inflammation in the ascending aorta and bilateral carotids were assessed using mean uptake of FDG as measured by PET. Regions of interest were drawn manually on anatomical MRI and translated to fused PET images of the Standard Uptake Value (SUV) – FDG uptake [Bq/ml] normalized by the injected dose, radioactive decay since injection, and body weight – and the mean SUV within the region of interest computed. More details can be found in the Supplemental Information.

### Imaging metrics and statistical methods

#### Brain connectivity measures

Network level connectivity between the amygdalae and the rest of the brain was calculated using Pearsons’s correlation for the fMRI timeseries and tractography counts for the dMRI data. More details can be found in the Supplemental Information.

#### Carotid artery analysis

Atherosclerotic burden was assessed using the average of the standard deviation of the wall thickness of each of the left and right common carotids (SDWT-C), as measured with black blood vessel wall MRI ^24,25^. More details can be found in the Supplemental Information.

#### Classification modelling

Logistic regression was performed, where the independent variables were connectivity estimates from brain MRI, and the dependent variable was SDWT of the bilateral carotids. Tenfold CV was used to avoid overfitting. To account for variation in train-test split, logistic regression was performed 1000 times. The median performing set of models was reported in this manuscript. To evaluate model performance, prediction accuracy was used. Models were also assessed using the receiver operating characteristic (ROC), and area under the curve (ROC-AUC). Permutation tests were performed to assess statistical significance of model performance. This was accomplished by shuffling the data with respect to participant labels 1000 times. Significance was then assessed based on the number of times actual model performance exceeded permuted model performance, divided by the number of iterations, giving an empirical p value.

#### Composite connectivity scores from classification models

For the successful models we used the median model weights to generate a summed connectivity network score. This was done by calculating the dot product of the median model weights across CV folds, for the median performing model, and the underlying connectivity estimates for each participant. This yielded one composite score per image, per participant. In each case the score reflects a weighted average of the connectivity between the amygdala and the 20 lateralized brain networks from the Shen atlas, which is sensitive to atherosclerosis (as per SDWT-C).

#### Association analyses

Composite scores were assessed for association with inflammation, self-report measures of stress and resilience, and brain PET. Ordinary least squares (OLS) regression was performed with correction for age, sex and participant motion in the scanner (mean framewise displacement, mean FD).

#### PET analysis

Prior work on the relationship between atherosclerosis, and SNA in PET imaging focused on using FDG uptake in the amygdala normalized by that of the vmPFC. The effect of normalizing the amygdala FDG uptake with distinct brain regions beyond the vmPFC was examined in this study, including sensorimotor and cerebellar regions. More details can be found in the Supplemental Information.

## Data Availability

Data will be made available upon reasonable request.

## Funding

This work is supported by NHLBI P01HL131478. SA is supported by American Heart Association (AHA) Second Century Early Faculty Independence Award 10.58275/AHA.24SCEFIA1256969.pc.gr.193937. MTO is supported by National Institutes of Health K23HL151909, American Heart Association 10.58275/AHA.23SCISA1143491.pc.gr.172152, and the generosity of the Hassenfeld family, MTO.

### Author contributions

Conceptualization: AT, ZAF

Methodology: DOC, PMR, TA, ZAF

Formal analysis: DOC, CG, SA

Visualization: DOC

Funding acquisition: AT, ZAF

Project administration: AT, ZAF

Supervision: AT, ZAF

Writing – original draft: DOC, MMTvL, LS, AT, ZF

Writing – review & editing: DOC, MMTvL, PMR, SA, CG, AEK, MTO, MGT, JWM, LMS, AT, ZAF

## Data and materials availability

Data will be made available upon reasonable request.

