## Supplemental Information for "Cortico-Limbic and Sensorimotor Network Connectivity Link Brain Function to Atherosclerosis in Chronic Stress: Beyond Amygdala-PFC"

### Dataset and screening

In this cohort 27 of the individuals had PTSD, 45 were exposed to a traumatic event but did not have PTSD (“Trauma Controls”, or TC), and 22 had neither trauma exposure nor PTSD (“Healthy Controls”, or HC). Diagnoses (or the lack thereof) were determined by which participants met the Diagnostic and Statistical Manual of Mental Disorders, 5^th^ edition (DSM-5) criteria for PTSD at the time of study enrollment. A Criterion A traumatic event happened at least one year before study enrollment. Participants in the TC group met the DSM-5 Criterion A traumatic event from at least one year before enrollment but did not meet other diagnostic criteria for PTSD. HC did not have any exposure to major traumatic events and did not meet the criteria for any current or past psychiatric diagnoses as defined by DSM-5. Psychiatric assessments were performed by experienced psychologists.

Participants were excluded if they had a clinical history of atherosclerotic CVD, chronic inflammatory conditions (e.g., rheumatoid arthritis, psoriasis), significant central nervous system or neurological disease, active malignancy, or malignancy within 5 years of remission, or a current primary psychiatric disorder other than PTSD. Participants were also excluded if they were on high-intensity statin therapy or select anti-inflammatory agents. A full list of inclusion and exclusion criteria is provided in the Supplement of Gharios et al ^1^.

Eligible participants underwent psychological evaluation, physical examination, blood draws, and FDG PET and MRI imaging. In accordance with previous work, 10 participants with hsCRP > 10 mg/L on the day of imaging were considered to likely have an acute inflammatory process and were excluded from analyses ^2,3^. The Icahn School of Medicine at Mount Sinai and the Mass General Brigham Institutional Review Boards approved the current study (17-00168 and 2017P000950, respectively), and all included participants provided informed consent.

### Image acquisition and preprocessing

#### **Structural MRI (sMRI)**

A 3D T1-weighted MPRAGE sequence was used to acquire a high-resolution anatomical scan of the brain. The parameters included: Inversion time (TI) of 900 ms, echo time (TE) of 2.49 ms, repetition time (TR) of 1900 ms, a flip angle (FA) of 9 degrees, voxel size of 0.9 mm x 0.9 mm x 1 mm, a field of view of 230 mm, 176 slices, an in-plane image matrix of 256x256, a slice thickness of 1 mm, phase encoding direction A >> P, and an acceleration factor of 2 with GRAPPA.

The T1-weighted (T1w) image was corrected for intensity non-uniformity, and skull-stripped. Brain tissue segmentation was performed to generate masks of grey matter, white matter, and cerebral spinal fluid tissues. The T1w images were then normalized to MNI space. Processing was performed using FSL ^4^, ANTs ^5^ and Freesurfer ^6^.

T1-weighted anatomical images were processed using the fMRIPrep 22.0.2 pipeline ^7^, which is built on Nipype 1.8.5 ^8^. Intensity non-uniformity was corrected using N4BiasFieldCorrection ^9^, and skull stripping was performed using the antsBrainExtraction.sh script with the OASIS template. Tissue segmentation into cerebrospinal fluid (CSF), white matter (WM), and gray matter (GM) was performed using FSL’s FAST ^10^. Nonlinear spatial normalization was performed using ANTs 2.3.3 (antsRegistration) to standard MNI spaces (ICBM152 nonlinear asymmetrical 2009c and 2006 templates) with reference templates sourced via TemplateFlow ^11^. Cortical surface reconstruction was carried out using FreeSurfer 7.1.1 (recon-all; ^12^), and anatomical segmentations from ANTs and FreeSurfer were reconciled using Mindboggle ^13^.

#### **Functional MRI (fMRI)**

Both rs-fMRI and t-fMRI data were acquired using the following parameters; TE of 27 ms, TR of 1900 ms, FA of 80 degrees, voxel size of 3 mm x 3 mm x 2.5 mm, a field of view of 220 mm, 50 slices, an in-plane matrix of 74x74, a slice thickness of 1 mm, phase encoding direction A >> P, and an acceleration factor of 2 with GRAPPA. The t-fMRI duration was 6 min, 56 sec with 266 volumes. The rs-fMRI was 10 min, with 400 volumes. The t-fMRI paradigm was the Hariri face-matching task, ^14,15^.

Functional images were also preprocessed using fMRIPrep 22.0.2 pipeline ^7^, which is built on Nipype 1.8.5 ^8^, and utilizes commands from FSL ^4^, AFNI^16^, Freesurfer ^12^ and ANTs ^5^. A skull-stripped BOLD reference image was generated, and susceptibility-induced distortions were estimated and corrected using a fieldmap derived from a gradient-recalled echo (GRE) sequence. Phase unwrapping was performed using FSL’s prelude. Head motion correction was conducted using FSL’s mcflirt, and slice timing correction was applied with AFNI’s 3dTshift. Co-registration of BOLD images to the T1-weighted anatomical space was performed using FreeSurfer’s bbregister, which employs boundary-based registration. The resulting images were normalized to MNI152NLin2009cAsym space using a composite transform that included motion correction, distortion correction, BOLD-to-T1w registration, and T1w-to-MNI normalization.

Confound regressors included framewise displacement (FD) computed using both Power et al. and Jenkinson methods, and global signal averages from CSF, WM, and whole-brain masks, and were applied using NiLearn ^17^. Final resampling of the BOLD time series was performed using ANTs’ antsApplyTransforms. A single interpolation step was used to minimize unnecessary smoothing.

#### **Diffusion MRI (dMRI)**

Two opposite phase-encoded (AP, PA) acquisitions using a NODDI sequence were obtained for the diffusion weighted MRI of the brain. For each the parameters were identical except the phase-encode direction, and included: A b-value of 2400 s/mm^2^, 107 directions, TE of 96.8 ms, TR of 4284 ms, FA of 90 degrees, with a refocusing FA of 180 degrees, voxel size of 2-mm isotropic, a field of view of 256 mm, 64 slices, an in-plane matrix of 128x128, a slice thickness of 2 mm, and a multiband acceleration factor of 2.

The dMRI data was preprocessed using the QSIPrep workflow ^18^. In short, dMRI data were denoised, corrected for B1 field inhomogeneity, eddy currents, motion, and susceptibility artifacts. dMRI were then normalized to MNI space. Multi-tissue fiber response functions were estimated using the dhollander algorithm. Fiber orientation distributions (FODs) were estimated via constrained spherical deconvolution (CSD)  ^19,20^, using an unsupervised multi-tissue method ^21,22^. Reconstruction was done using MRtrix3 ^23^. FODs were intensity-normalized using mtnormalize ^24^.

Diffusion-weighted images were processed using MRtrix3, FSL, and ANTs through the QSIPrep pipeline (version 0.16.1; based on Nipype 1.8.5; ^8,18,25^). Thermal noise was removed using the MP-PCA algorithm implemented in MRtrix3’s dwidenoise ^26^, followed by bias field correction using dwibiascorrect with the N4 algorithm. Correction for head motion, eddy currents, and susceptibility-induced distortions was performed using FSL’s eddy, including outlier replacement and intra-volume motion modeling ^27^. Distortion correction was guided by reverse phase-encoded images used to estimate the susceptibility-induced off-resonance field. The mean b0 image was aligned to the AC–PC line using a rigid-body transformation, and final diffusion data were resampled to 2 mm isotropic resolution. Framewise displacement was computed across the diffusion time series, and slice-wise signal dropout was quantified via Pearson correlation between corresponding slices across volumes.

QSIPrep-preprocessed T1-weighted images and brain masks were used to guide diffusion reconstruction. A hybrid surface/volume segmentation was created to facilitate anatomically-informed processing ^28^, and FreeSurfer outputs were registered to the QSIPrep anatomical space. Fiber orientation distributions (FODs) were reconstructed using MRtrix3 ^23^. Multi-tissue fiber response functions were estimated with the unsupervised Dhollander algorithm, and FODs were computed using constrained spherical deconvolution (CSD; ^19,20^) via a multi-tissue approach ^21,22^. FODs were intensity-normalized using the mtnormalize algorithm ^24^. QSIPrep internally employs Nilearn 0.9.2 ^17^ and DIPY 1.5.0 ^29^ for image processing and tractography. Further pipeline details are available in the QSIPrep workflow documentation.

#### Vascular and Leukopoietic MRI and PET

3-D dark-blood MRI imaging of the internal carotid arteries and ascending aorta was performed. Carotid imaging extending 3 cm below and above the carotid bifurcations was conducted using a 3-D SPACE acquisition. Separate images with proton density, T1, and T2 weighting were acquired. Acquisition parameters included field of view 200x175 mm^2^, number of slices 64, isotropic resolution 0.8x0.8x0.8 mm^3^, fat suppression, acceleration factor 2 (GRAPPA). Weighting-specific parameters included TR 2000/650/2000 ms, TE 25/21/135 ms, turbo factor 117/59/129, vendor-determined echo train optimization PDvar/T1var/T2var, averages 1.4/2/2, phase over-sampling 95%/100%/4%, acquisition time 6min 42sec/6min 17sec/4min 50sec, respectively.

Measurement of vascular and leukopoietic tissue activity and structure was conducted using MRI- and PET-based approaches. MRI-derived parameters for assessing atherosclerotic burden at the carotid arteries and ascending aorta included lumen area, wall area, total vessel area, wall thickness, and the standard deviation of wall thickness. The normalized wall index (NWI) was calculated as the ratio of wall area to total vessel area. For carotid arteries, measurements were averaged across the left and right sides.

Arterial 18F-FDG uptake, representing atherosclerotic inflammation, was quantified at the bilateral common carotid arteries and the ascending aorta using maximal standardized uptake values (SUV) The carotid SUVmax values were averaged bilaterally. To derive the target-to-background ratio (TBR), venous background activity was measured as the average mean SUV within the lumen of the bilateral jugular veins (for carotids) and the superior vena cava (for aorta) in regions free of significant spillover. TBRmax was computed as the average maximum arterial SUV divided by the background venous mean SUV ^30,31^.

Leukopoietic tissue 18F-FDG uptake, a marker of metabolic activity and leukopoiesis, was assessed in the spleen and bone marrow (cervical through lumbar vertebral bodies). TBRmax for spleen and bone marrow was calculated using venous background from the superior vena cava.

### Statistical Analysis

#### Functional Connectivity

The Shen 368 atlas was used to parcellate both fMRI and dMRI. The Shen 368 atlas includes cortical regions of interest generated from clustering resting state fMRI ^32^, anatomic delineation of subcortical regions, and a cerebellum parcellation based on the Yeo 17-network parcellation ^33^. Parcellation of the fMRI entailed calculating the mean blood oxygenation level dependent (BOLD) signal for each region in the atlas and then estimating the pairwise temporal correlation of all regions in the atlas resulting in a symmetric 368 x 368 matrix. In the case of dMRI this involved counting the number of streamlines from the tractography results between each region in the atlas, also resulting in a symmetric 368 x 368 matrix. In each case data were pared down to connections between each amygdala and the rest of the brain only. The region level estimates were averaged into hemispheric and network-wise mean connections, e.g. the mean connectivity between the amygdala and right default mode network and between the amygdala and the left frontoparietal network. This was done for the right amygdala (omitting the left) and left amygdala (omitting the right). This resulted in a vector of length 20 for each amygdala, which represents the connectivity between the amygdala and all 10 networks split across the two hemispheres. For bilateral amygdalae assessment these results were concatenated, resulting in a vector of length 40.

#### Carotid Artery Analysis

Briefly, this method dephases the signal from flowing blood in the lumen in the vessel of interest, making the lumen dark, allowing for optimal image contrast with the vessel wall. This makes detection of structural abnormalities easier, including plaque formation and vessel wall remodeling. Plaque formation results in either lumen narrowing or positive remodeling, and in either case uneven widening of the vessel wall circumferentially. This can be captured by measuring the circumferential variability of the vessel wall as SDWT-C.

#### Brain PET Image Analysis

This was accomplished using the pyapetnet reconstruction algorithm ^34^, the Shen 368 atlas, as described in “Brain connectivity measures”, and custom python scripts. Pyapetnet is a convolutional neural network designed to do anatomically guided reconstruction of PET imaging. Specifically, the algorithm performs PET reconstruction based on the asymmetric Bowsher prior. Shramm et al. show that the Bowsher prior is well-approximated by a purely shift-invariant convolutional neural network in image space, allowing the generation of anatomically guided PET images. A biproduct of this reconstruction is a PET image that is very well aligned with the participants T1 anatomical image. This allows for the PET data to be moved into a common space via the non-linear registration of the T1 image, and the subsequent parcellation of the PET using a common space atlas, in this case the Shen 368 atlas. Amygdala FDG uptake normalized by each of the 368 Shen atlas regions was then calculated, and related to SDWT-C using OLS, accounting for age and sex. In this analysis adjusted t-values are reported. Multiple comparisons correction was not performed.

### Supplemental Results

#### Classification metrics

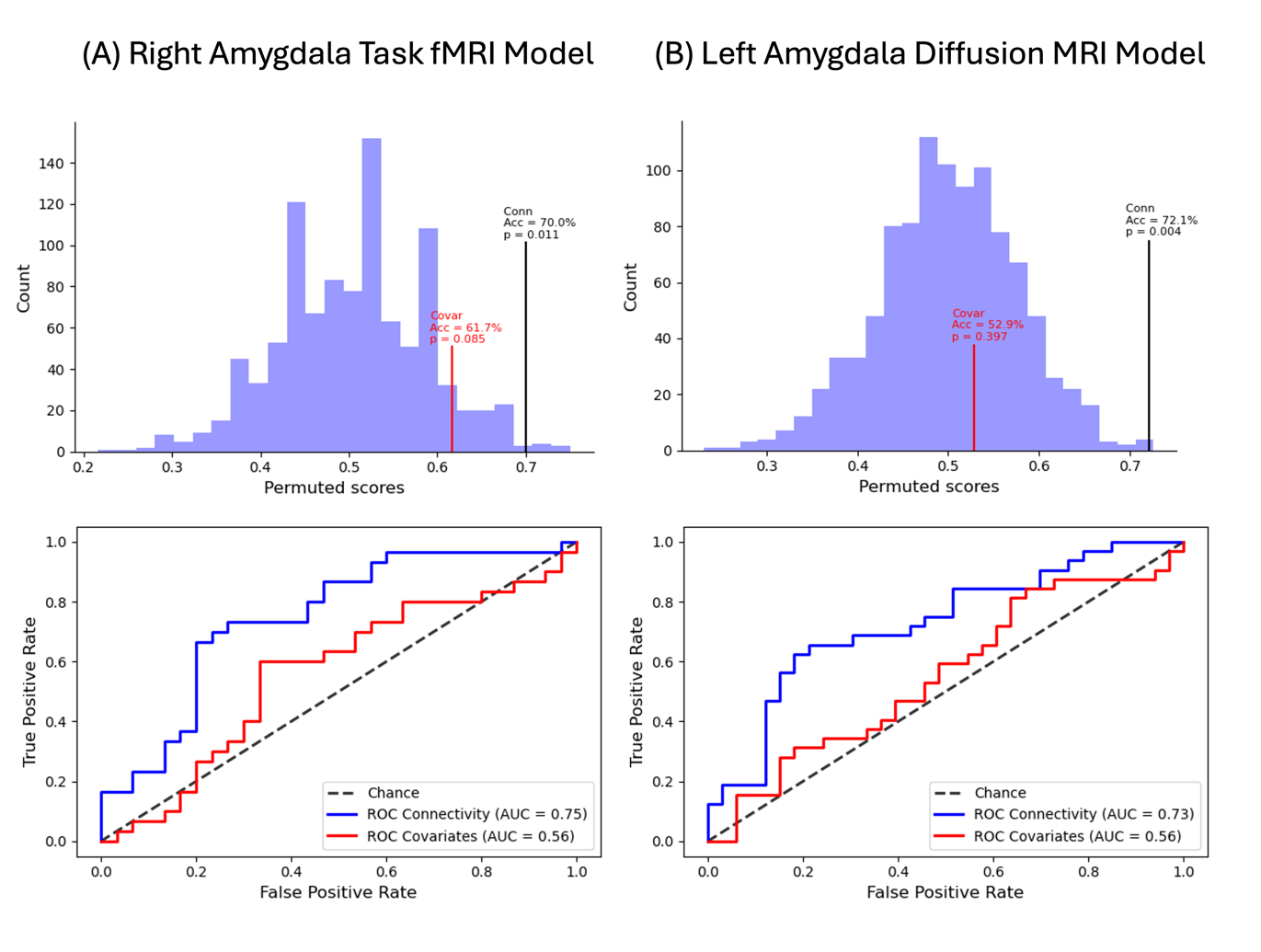

Figure 1 – Performance of successful functional and structural connectivity models. **Column A** shows the permutation distribution and p values for the successful task fMRI model (top row) and the ROC-AUC assessment (bottom row). **Column B** shows the permutation distribution and p values for the successful dMRI model (top row) and the ROC-AUC assessment (bottom row). The effect of predicting with covariates of non-interest (age, sex and scanner motion) are also shown in red in (A) and (B). Connectivity scores outperform covariates in each case.

#### Regression models

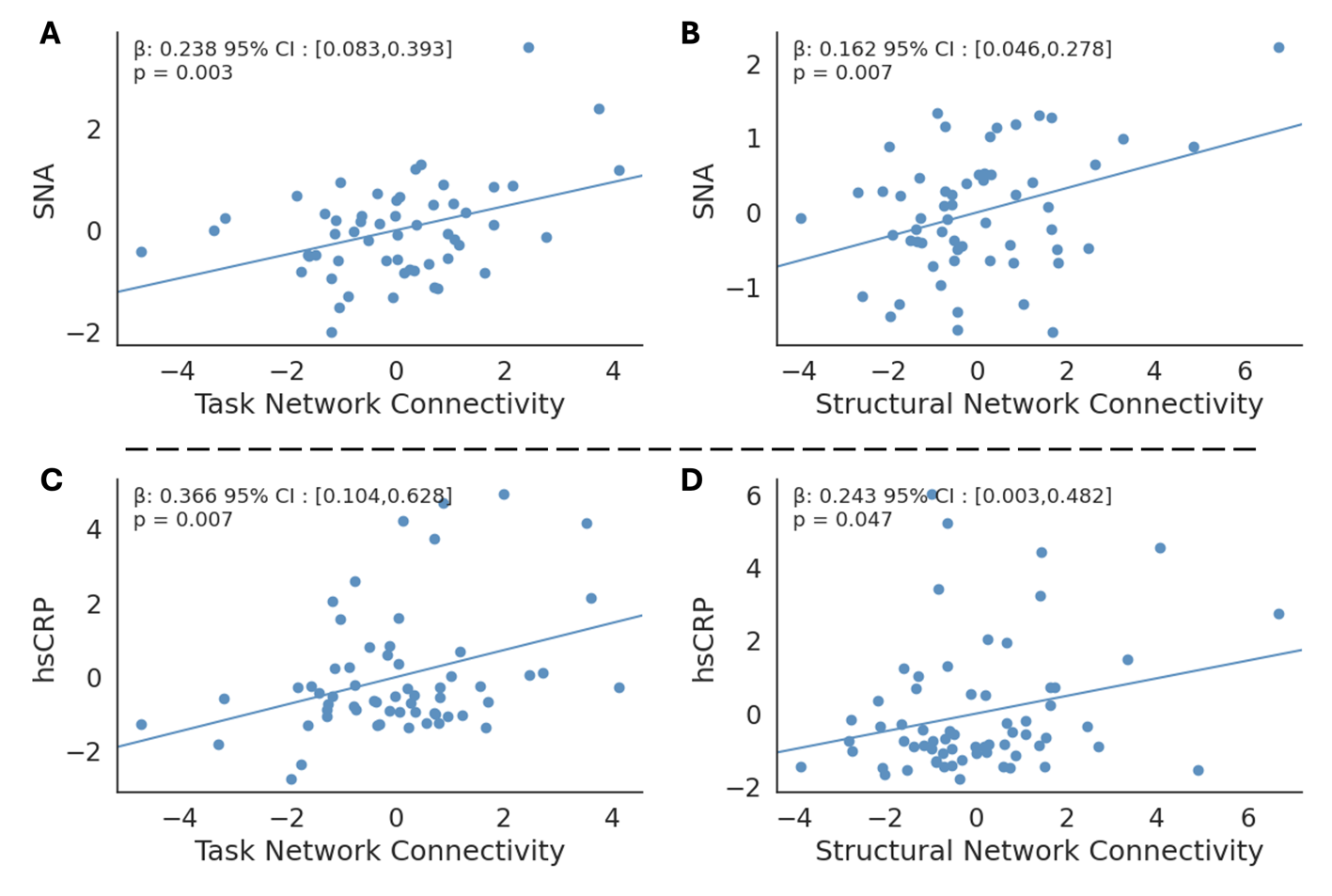

Figure 2 - Associations between composite connectivity scores and stress related neural activity from PET (SNA), and systemic inflammation (high sensitivity CRP, hsCRP). **Panel A** shows the association between task-based functional connectivity composite score and SNA. **Panel B** shows the association between the structural connectivity composite score and SNA. **Panel C** shows the association between task-based functional connectivity composite score and hsCRP. **Panel D** shows the association between the structural connectivity composite score and hsCRP. Statistical parameters were generated using OLS, and are corrected for age, sex and in scanner participant motion.

Models were run using age, sex and in scanner motion as covariates. Reported are the statistics for the variable of interest only.

Table 1 – Regression results for network composite scores and non-brain MRI metrics.

| Modality | Outcome | Beta | 95% CI | t | Uncorrected p value | N |
| --- | --- | --- | --- | --- | --- | --- |
| Task fMRI | PSS | 0.273 | [-1.082,1.628] | 0.404 | 0.688 | 60 |
| Task fMRI | STICSA | 0.121 | [-1.424,1.667] | 0.157 | 0.875 | 60 |
| Task fMRI | CDRISC | 0.711 | [-1.865,3.288] | 0.553 | 0.582 | 60 |
| Task fMRI | PCL | 0.545 | [-3.795,4.884] | 0.257 | 0.799 | 34 |
| Task fMRI | hsCRP | 0.366 | [0.104,0.628] | 2.801 | 0.007 | 60 |
| Task fMRI | Spleen TBR Max | 0.015 | [-0.013,0.043] | 1.066 | 0.291 | 57 |
| Task fMRI | Marrow TBR Max | 0.028 | [-0.005,0.061] | 1.729 | 0.09 | 57 |
| Task fMRI | Carotid TBR Max | -0.025 | [-0.16,0.111] | -0.366 | 0.716 | 49 |
| Task fMRI | Aorta TBR Max | 0.044 | [-0.101,0.19] | 0.614 | 0.542 | 56 |
| Task fMRI | Aorta NWI | 0.018 | [-0.004,0.041] | 1.641 | 0.108 | 51 |
| Task fMRI | SNA | 0.238 | [0.083,0.393] | 3.082 | 0.003 | 55 |
| dMRI | PSS | -0.178 | [-1.267,0.911] | -0.327 | 0.745 | 65 |
| dMRI | STICSA | -0.188 | [-1.524,1.148] | -0.282 | 0.779 | 65 |
| dMRI | CDRISC | 1.373 | [-0.537,3.283] | 1.438 | 0.156 | 65 |
| dMRI | PCL | 0.972 | [-3.44,5.385] | 0.449 | 0.657 | 37 |
| dMRI | hsCRP | 0.243 | [0.003,0.482] | 2.027 | 0.047 | 65 |
| dMRI | Spleen TBR Max | 0.01 | [-0.014,0.033] | 0.827 | 0.412 | 63 |
| dMRI | Marrow TBR Max | 0.01 | [-0.02,0.039] | 0.652 | 0.517 | 63 |
| dMRI | Carotid TBR Max | -0.026 | [-0.149,0.098] | -0.417 | 0.678 | 52 |
| dMRI | Aorta TBR Max | -0.036 | [-0.156,0.084] | -0.599 | 0.551 | 61 |
| dMRI | Aorta NWI | -0.009 | [-0.029,0.011] | -0.915 | 0.364 | 54 |
| dMRI | SNA | 0.162 | [0.046,0.278] | 2.8 | 0.007 | 58 |

PCL-5: PTSD Checklist for DSM-5 (self-reported)

PSS: Perceived Stress Scale

CD-RISC: Connor-Davidson Resilience Scale

STICSA: State-Trait Inventory of Cognitive and Somatic Anxiety

hsCRP: High-sensitivity C-reactive protein

SNA: Stress-associated neural network activity

Table 2 –Functional MRI based network membership of regions shown in **Figure 5**. MF: medial frontal, FP: frontoparietal, DM: default mode, M: motor, VI: visual I, VII: visual II, VA: visual association, OC: cingulo-opercular cortex, C: cerebellum.

| Network | No. of Regions | Right Hemisphere | Left Hemisphere |
| --- | --- | --- | --- |
| MF | 9 | 4 | 5 |
| FP | 20 | 12 | 8 |
| DM | 6 | 1 | 5 |
| M | 3 | 2 | 1 |
| VI | 3 | 1 | 2 |
| VII | 10 | 4 | 6 |
| VA | 2 | 0 | 2 |
| OC | 6 | 2 | 4 |
| C | 2 | 0 | 2 |

1.	Gharios C, van Leent MMT, Chang HL, et al. Cortico-limbic Interactions and carotid atherosclerotic burden during chronic stress exposure. *European Heart Journal*. Published online 2024.

2.	Ridker PM. A Test in Context: High-Sensitivity C-Reactive Protein. *J Am Coll Cardiol*. 2016;67(6):712-723. doi:10.1016/j.jacc.2015.11.037

3.	Seo WW, Kim HL, Kim YJ, et al. Incremental prognostic value of high-sensitive C-reactive protein in patients undergoing coronary computed tomography angiography. *J Cardiol*. 2016;68(3):222-228. doi:10.1016/j.jjcc.2015.09.010

4.	Smith SM, Jenkinson M, Woolrich MW, et al. Advances in functional and structural MR image analysis and implementation as FSL. In: *NeuroImage*. Vol 23. Academic Press; 2004:S208-S219. doi:10.1016/j.neuroimage.2004.07.051

5.	Avants BB, Tustison N, Johnson H. Advanced Normalization Tools (ANTS). Published online July 10, 2014.

21.	Dhollander T, Raffelt D, Connelly A. *Unsupervised 3-Tissue Response Function Estimation from Single-Shell or Multi-Shell Diffusion MR Data without a Co-Registered T1 Image*.; 2016.

22.	Dhollander T, Mito R, Raffelt D, Connelly A. *Improved White Matter Response Function Estimation for 3-Tissue Constrained Spherical Deconvolution*.; 2019.
